# Effect of the COVID-19 pandemic on the post-Ebola referral system in Sierra Leone: A descriptive analysis

**DOI:** 10.1101/2025.03.05.25323450

**Authors:** Sorie Samura, Daniel Youkee, Bailah Molleh, Joseph Sam Kanu, Matthew Jusu Vandy, Songor Koedoyoma, Thaimu Mohamed Bangura, Hawa K. Lansana, Ibrahim Foday Musa, Mustapha S. Kabba, Sartie M. Kenneh, Adrienne K. Chan, Sharmistha Mishra, Stephen Sevalie, Sulaiman Lakoh

## Abstract

**Background:** The COVID-19 pandemic severely impacted health systems in several countries, including Sierra Leone, which was recovering from the 2014-2016 Ebola Virus Disease epidemic. The National Referral Service was established in 2017 followed by launching of the National Emergency Medical Services (NEMS) in 2018 to strengthen the country’s referral system as part of the post-Ebola recovery strategy. This study examined the impact of COVID-19 on the referral system and hospital bed occupancy in Sierra Leone.

**Methods:** We conducted a retrospective study using individual-level referral and daily hospital bed occupancy data from 16 public hospitals in Sierra Leone, covering the period October 2017 to February 2022. Data were categorized into four periods: post-Ebola/pre-NEMS (12 months), post-NEMS/pre-COVID (12 months), during COVID-19 (12 months), and post-COVID (12 months). Descriptive statistics was used to analyze the trends in referral and bed occupancy.

**Results:** A total of 78,406 referrals were recorded. Referrals increased from 14,870 in the post-Ebola/pre-NEMS period to 27,299 to post-NEMS launch. The number of referrals dropped to 22,948 (during COVID-19) and 13,289 (post-COVID-19). The mean number of referrals per facility initially increased from 1,359 in the post-Ebola and pre-NEMS periods but declined to 1,863 during and after the pandemic. The number of referrals at the provincial level was consistently higher than in Freetown. Bed occupancy rates also declined significantly during the COVID-19 pandemic, with a more pronounced decline in Freetown.

**Conclusion:** Our results show a continued decline in patient referrals and bed occupancy even after the peak of the COVID-19 pandemic, with greater impact in the capital Freetown than the provincial regions. The initial increase in referrals and bed occupancy post-Ebola reflects health system strengthening interventions following the Ebola outbreak. However, the COVID-19 pandemic disruptions led to significant declines in both metrics, underscoring the need for sustainable investment in resilient health systems.

## Background

Referral systems are a cornerstone of healthcare delivery. Functional referral systems enable flow of care from primary to secondary/tertiary health services and vice versa [1]. In Sierra Leone, the scale of the 2014-2016 Ebola Virus Disease epidemic exposed weaknesses in key aspects of the country’s health system, including the absence of a functional referral system and lack of coordination between healthcare providers and the general population [2-4]. This resulted in long delays in moving patients across the health system to access appropriate care, leading to higher mortality amongst all category of patients during the Ebola epidemic [5, 6].

As part of lessons learnt from the Ebola epidemic, the Ministry of Health of Sierra Leone established the National Referral Service in September 2017 [7]. The mandate of the National Referral Service was to strengthen patient referral pathways, map national healthcare services and create communication pathways between different levels of service delivery points and support hospital preparedness for emergency referrals [8]. Referrals are integrated into the configuration of the Sierra Leone public health system, which is divided into primary, secondary and tertiary levels. If a patient at a lower level service delivery point (e.g. a woman with obstetric complication at a community health centre) cannot be cared for effectively at this lower level, the patient is referred to a district secondary hospital using the National Referral System [9]. They also coordinate counter referrals from secondary or tertiary hospitals to peripheral health units for continuation of care after major medical or surgical intervention. This newly formed system leveraged an existing network of referral coordinators, which initially supported the provision of free health care to Ebola survivors during the 2014-2016 Ebola Virus Disease epidemic. The National Referral System consists of a network of referral coordinators with clinical background in nursing and community health and additional training in communication, data management and referrals skills. They are deployed in each district hospital to facilitate all incoming (from other facilities) and outgoing (from the district hospital to other facilities, such as tertiary hospitals) referrals. Following the operationalization of the National Referral System, the Ministry of Health then established the National Emergency Medical Services (NEMS) to provide free ambulance transportation services in October 2018 with interoperability with the National Referral Service System [9].

The joint operationalization of these two systems was put to test by the COVID-19 pandemic in Sierra Leone in 2020. COVID-19 preventive measures such as lockdown and movement restrictions to reduce the scale of the outbreak led to disruptions to health services in Sierra Leone, with an increase in non-COVID-19 related mortality [10-12].

Thus, we sought to describe the use of the National Referral System established in 2017 (following the 2014-2016 Ebola Virus Disease epidemic), and after the introduction of the ambulance system in 2018, up to and including 2022. We then sought to examine the effect of COVID-19 on hospital bed occupancy and the use of the National Referral System, stratified by districts, level of care, and categories of patients.

## Methods

### Study Design

The study involved a narrative description of the National Referral system and a retrospective analysis of a longitudinal cohort of individual-level referral data, and daily hospital bed occupancy data, both from the National Referral Coordination Unit of NEMS of the Ministry of Health.

### Study Population

The study utilized 78,406 individual patient referral data and daily hospital bed occupancy data from 16 public hospitals in Sierra Leone. Data for all referrals and bed occupancy data collected within the study period was included.

### Study Setting

#### General setting

Sierra Leone, a low-income country in West Africa with a population of approximately 7.8 million. It has five geographical regions, including the Western Area (urban and rural), Eastern, Northern, Southern and Northwestern regions (Figure 1) [13]. The country’s health system has faced significant challenges in recent years with its healthcare infrastructure severely weakened by a decade-long civil war that ended in 2002, and further strained by the devastating 2014-2016 Ebola Virus Disease epidemic [14]. These events have had long-lasting impacts on the country’s ability to provide quality healthcare to its citizens. This is further complicated by the weak national health financing system and high out-of-pocket payment for health services [15].

**Figure 1:**
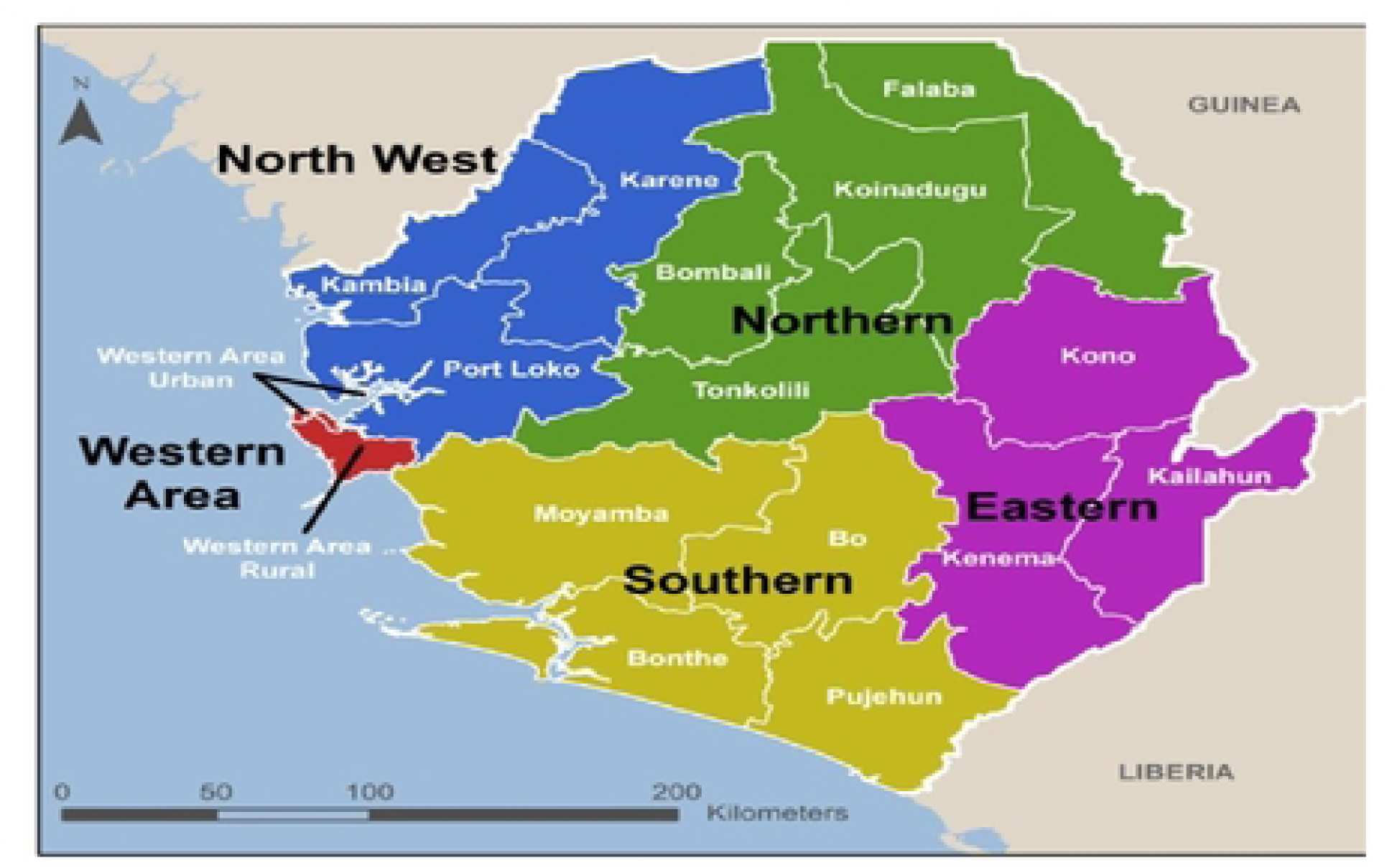
Administrative map of Sierra Leone with regions and districts indicated. Freetown is in the Western Area Urban district.

#### Specific setting

The public health system in Sierra Leone is divided into primary, secondary and tertiary care. Primary health facilities consist of Maternal and Child Health Posts, Community Health Posts and Community Health Centers, which provide basic emergency obstetric and newborn care and treat common febrile illness. The secondary level facilities consist of district hospitals, whilst the tertiary level consist of national referral/regional hospitals. These two levels provide Comprehensive Emergency Obstetric and Newborn Care services and adult surgical and medical services. There are 35 public hospitals in Sierra Leone.

To support patient referrals to these hospitals, the Government of Sierra Leone designed a referral coordination system in 2016. The national referral coordination system was launched in September 2017 with referral coordinators posted to all secondary and tertiary hospitals to support referrals for all categories of patients across the country. Following this, the Ministry of Health launched the NEMS to provide free ambulance transportation services to all patients with emergency conditions to strengthen the referral system. These two systems collectively coordinate referrals of all patients across all levels of the health care system. NEMS has a network of 102 ambulances as of September 2024 with trained paramedics that are geospatially distributed across all 16 districts. The movement of these ambulances is coordinated by a central operation system in the capital Freetown (accessible to citizens through a toll-free line).

There are 52 referral coordinators in 22 government facilities that coordinate all referrals, support hospital preparedness by ensuring that beds and services are available, and clinicians are in readiness to receive incoming patient and collect data and provide feedback on all referred patient.

Referrals are initiated with a telephone call from clinician in any health facilities requesting for an ambulance for a patient that cannot be managed at that level. These calls are triaged by the NEMS operators who assign ambulance based on severity of the case, service availability in the nearest hospital as confirmed through the referral coordinators in the receiving facility. The referral coordinators then use the information provided by the referring clinician and the NEMS operators to prepare the hospital for the incoming patient. Flow chart of referral process shown in annex 1.

Overall, the National Referral Services aims to enhance access and navigation to services and improve hospital preparedness, patient flow, systemic coordination and information feedback (Figure 2).

**Figure 2:**
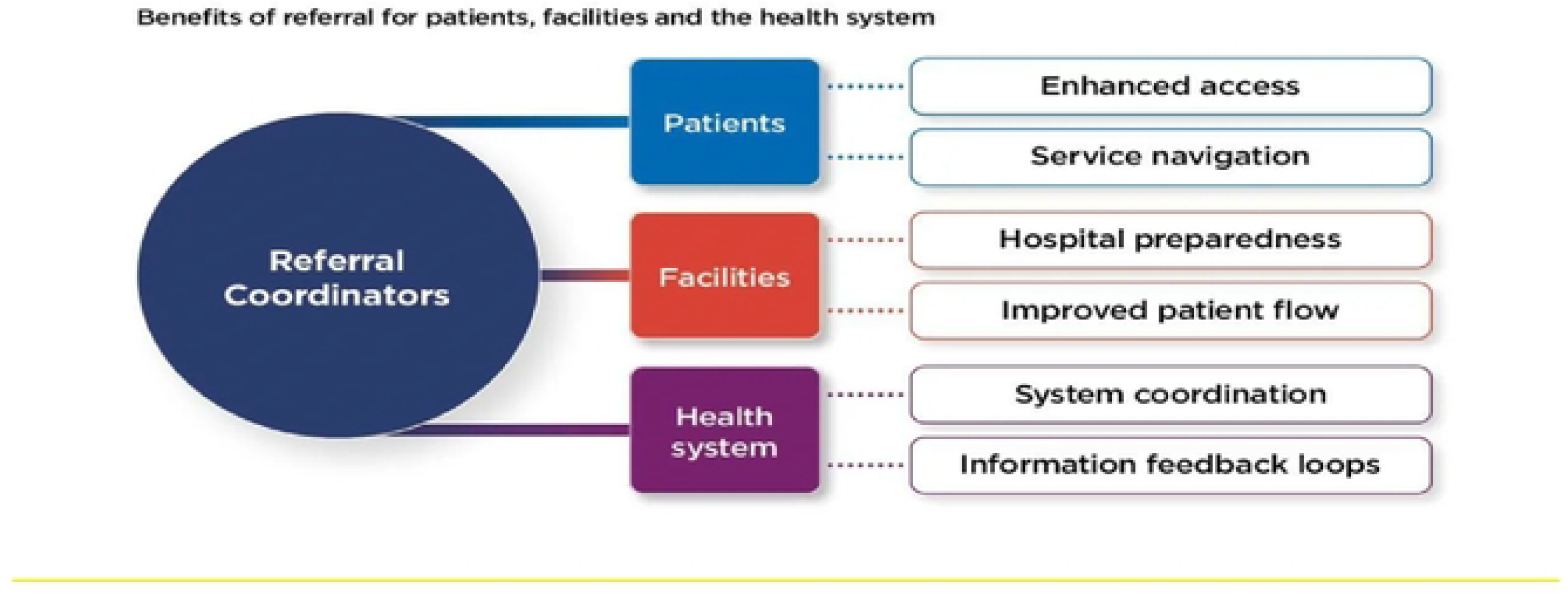
Aims of the referral network.

Data from 18 hospitals were included in this study. Generally, these hospitals have bed capacity ranging between 30 and 300 beds. The technical leadership for referrals at each hospital is provided by the Referral Coordinators who collect data.

### The Impact of Ebola Outbreak on Sierra Leone Health System

The 2014-2016 Ebola Virus Disease epidemic had a profound impact on Sierra Leone’s health system, which resulted in the deaths of nearly 4,000 people, including many healthcare workers and severely weakened the healthcare workforce and infrastructure. The outbreak exposed significant weaknesses in infection prevention and control practices, surveillance, laboratory capacity, and the referral system [2]. Healthcare utilization plummeted during the outbreak due to fear of infection and the collapse of health services. For instance, there were marked reductions in pediatric and maternity admissions, as well as in the utilization of maternal and child health services [16,17]. This led to increased all-cause mortality, particularly among children under five, and a rise in preventable diseases due to disruptions in vaccination programs [2]. The epidemic also strained the health system’s leadership and governance, disrupting health programs and services. Efforts to rebuild the health system post-Ebola focused on strengthening laboratory infrastructure, emergency medical services, and referral systems. However, the pandemic highlighted the need for sustained international support to fully restore and enhance Sierra Leone’s health system resilience [2, 16].

### The impact of Covid-19 on Sierra Leone Health System

The COVID-19 pandemic has had profound impacts on Sierra Leone’s health system. Whilst the country is still recovering from the 2014-2016 Ebola Virus Disease epidemic, faced significant disruptions in healthcare delivery due to the pandemic [18]. One of the most notable impacts was the decline in utilization of essential health services [18, 19]. Studies have documented substantial reductions in outpatient visits, maternal and child health services, and routine immunizations [20].

Healthcare workers in Sierra Leone faced increased stress, anxiety, and burnout due to the pandemic. The added workload, coupled with inadequate protective equipment and fear of infection, significantly impacted their ability to provide quality care [21].

Despite these challenges, Sierra Leone’s health system demonstrated some resilience. The country leveraged lessons and systems established during the 2014-2016 Ebola Virus Disease epidemic to respond to COVID-19. This included activating emergency operations centers, deploying rapid response teams, and utilizing community health workers for surveillance and contact tracing [22]. Telemedicine initiatives were also expanded to overcome referral challenges, showing promise in reducing unnecessary referrals and improving care coordination [18].

### Study period

Data was analysed from all the referrals and hospital bed occupancy from October 2017 to February 2022 over four time periods; post-Ebola and before the establishment of NEMS (October 2017 to November 2018), after the establishment of NEMS and before Covid-19 (December 2018 to February 2019), during COVID-19 (March 2020 to February 2021) and post COVID-19 (March 2021 to February 2022) using secondary referral and bed occupancy data (Figure 3).

**Figure 3:**
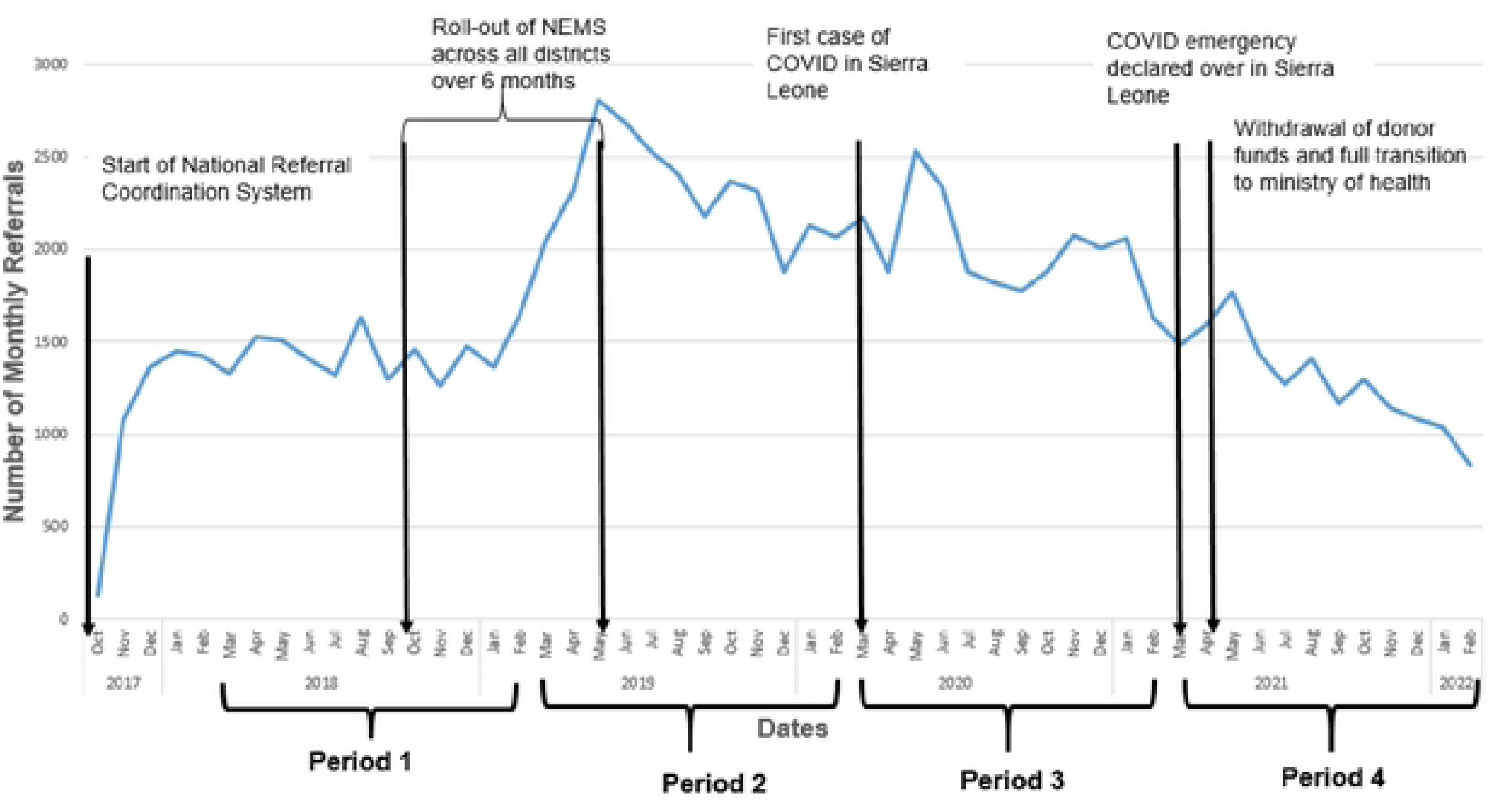
Time periods of the study and key events. *NFMS: National Emergency Medical Services*

### Data Sources

Data for this study was extracted on September 20, 2023 from the National Referral Coordinator’s database consisting of secondary, de-identified, individual patients and hospital bed occupancy data collected daily at government secondary and tertiary hospitals. The referral dataset is recorded by the Referral Coordinators in all public hospitals across the country. The data for all referred patients is recorded on paper forms by each hospital’s referral coordinator, then transferred to an electronic EpiInfo™ data sheet, which is then exported to an excel sheet.

The bed occupancy data is reported daily by Referral Coordinators via SMS/WhatsApp and compiled via excel with quality assurance provided by the central referral support team in Freetown.

### Analysis

We visualized trends in monthly referrals and timing of key events to define the study periods. We conducted descriptive statistics and reported the median and interquartile range of monthly referrals, by region (Western Area versus provincial). To estimate the association between time-period and referrals (count outcome), we conducted negative binomial regression (given overdispersion). We reported the incidence rate ratio and 95% confidence intervals (CIs), using the first period (post-Ebola) as the reference group. Finally, we described and visualized monthly bed occupancy over time, and by region (Western Area versus provincial). We used MS Excel to generate figures and used Stata for the regression analysis.

### Ethics

Ethics approval was obtained from the Sierra Leone Ethics and Scientific Review Committee of the Ministry of Health, Government of Sierra Leone with ethical approval number 020/05/2023. The study involved secondary analysis of anonymised service level data, abstracted under a waiver of informed consent from the ethics committee with the approval of NEMS. The authors did not have access to information that could identify individual participants during or after data collection.

## RESULTS

### Referrals across districts in Sierra Leone

Between March 2018 and February 2022, we recorded 78, 406 referrals across 16 public hospitals. There were 14,870 referrals in the pre-NEMS and post-Ebola period. The number of referrals increased to 27,299 after launching the national ambulance services, but reduced to 22,948 in the COVID-19 period and 13,289 in the post-COVID-19 period. Of all the referrals, 24,375 were in the capital Freetown (Table 1). The median national referrals per facility increased from 1,359 in the post-Ebola and pre-NEMS period to 2,320 in the post-Ebola but declined to 1,863 during COVID-19. The drop in median referrals in provincial regions was greater than the drop in median referrals in the capital Freetown (Table 2a). There is higher incident rate ratio in provincial regions than Freetown, across all time periods, and a larger decrease in incident rate ratio in F|reetown during COVID-19 period compared to provincial regions(Table 2b).

**Table 1:**
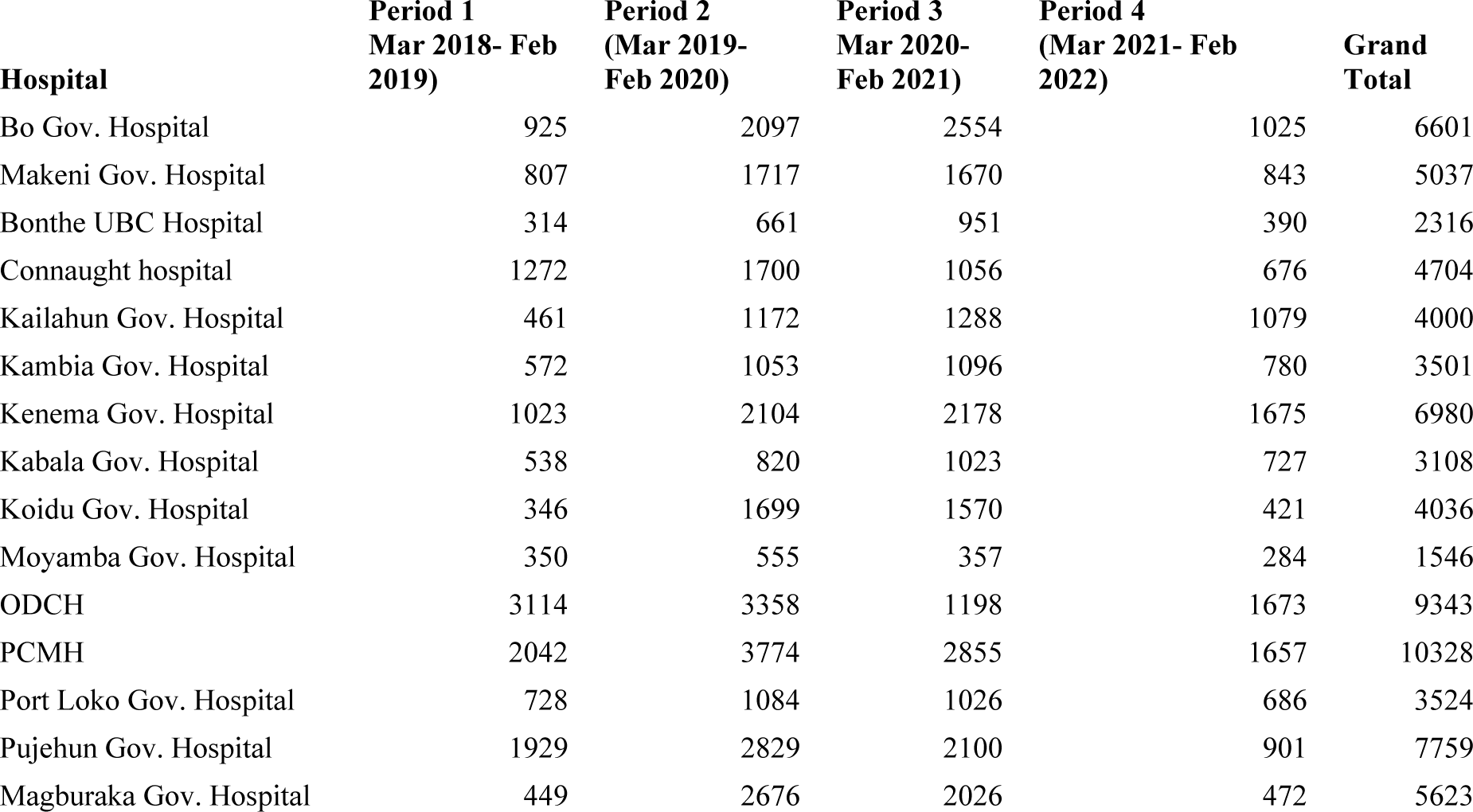
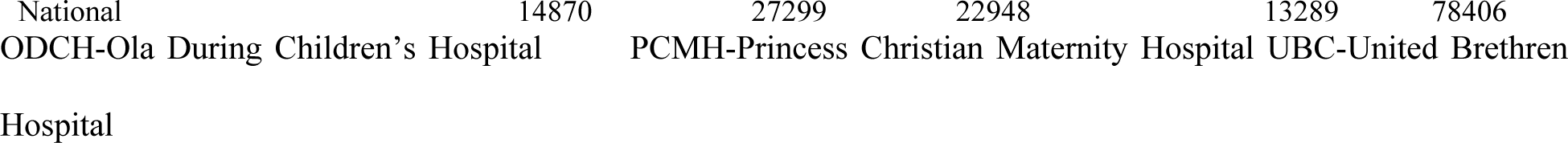
Total number of referrals across different time periods and hospitals.

| Hospital | Period 1<br>Mar 2018- Feb<br>2019) | Period 2<br>(Mar 2019-Feb 2020) | Period 3<br>Mar 2020-Feb 2021) | Period 4<br>(Mar 2021- Feb<br>2022) | Grand<br>Total |
| --- | --- | --- | --- | --- | --- |
| Bo Gov. Hospital | 925 | 2097 | 2554 | 1025 | 6601 |
| Makeni Gov. Hospital | 807 | 1717 | 1670 | 843 | 5037 |
| Bonthe UBC Hospital | 314 | 661 | 951 | 390 | 2316 |
| Connaught hospital | 1272 | 1700 | 1056 | 676 | 4704 |
| Kailahun Gov. Hospital | 461 | 1172 | 1288 | 1079 | 4000 |
| Kambia Gov. Hospital | 572 | 1053 | 1096 | 780 | 3501 |
| Kenema Gov. Hospital | 1023 | 2104 | 2178 | 1675 | 6980 |
| Kabala Gov. Hospital | 538 | 820 | 1023 | 727 | 3108 |
| Koidu Gov. Hospital | 346 | 1699 | 1570 | 421 | 4036 |
| Moyamba Gov. Hospital | 350 | 555 | 357 | 284 | 1546 |
| ODCH | 3114 | 3358 | 1198 | 1673 | 9343 |
| PCMH | 2042 | 3774 | 2855 | 1657 | 10328 |
| Port Loko Gov. Hospital | 728 | 1084 | 1026 | 686 | 3524 |
| Pujehun Gov. Hospital | 1929 | 2829 | 2100 | 901 | 7759 |
| Magburaka Gov. Hospital | 449 | 2676 | 2026 | 472 | 5623 |

**Table 2a:** Median number of referrals across periods of COVID-19 between March 2018 and February 2022 Referral categories Study periods.

| Referral categories | Study periods |  |  |  |
| --- | --- | --- | --- | --- |
|  | Post-Ebola<br>(March 2018 to<br>February 2019)<br>Median (IQR) | Pre-COVID-19<br>(March 2019 to<br>February 2020)<br>Median (IQR) | COVID-19<br>(March 2020 to<br>February 2021)<br>Median (IQR) | Post-COVID-19<br>(March 2021 to<br>February 2022)<br>Median (IQR) |
| National | 1359<br>(1226 – 1453) | 2320<br>(2081 – 2502) | 1863<br>(1779 – 2127) | 1121<br>(992 – 1271) |
| Freetown | 612<br>(495 – 691) | 715<br>(667 – 813) | 416<br>(363 – 443) | 323<br>(295 – 372) |
| Provincial | 763<br>(687 – 818) | 1530<br>(1438 – 1783) | 1461<br>(1370 – 1581) | 811<br>(846 – 899) |
| Under 5s | 556<br>(495 – 661) | 758<br>(618 – 1005) | 477<br>(408 – 633) | 328<br>(278 – 295) |
| Obstetric | 651<br>(586 – 700) | 1007<br>(959 – 1071) | 1032<br>(913 – 1066) | 578<br>(472 – 646) |
| Ebola Survivors | 23<br>(19 – 30) | 0 (0) | 0 (0) | 0 (0) |
| Non-FHCI | 140<br>(121 – 153) | 457<br>(418 – 475) | 369<br>(345 – 409) | 177<br>(149 – 215) |
| Disabled/Destitute | 11(9 – 11) | 8(6 – 9) | 8(6 – 25) | 4(1 – 5) |

FHCI-Free Healthcare Initiative IQR-Interquartile range

**Table 2b:** Trends in monthly referrals and the effect of the COVID-19 pandemic on the referral system.

| Referral categories | Study periods |  |  |  |
| --- | --- | --- | --- | --- |
|  | Post-Ebola | Pre-COVID-19 | During COVID-19 | Post-COVID-19 |
|  | Ref | IRR (95% CI) | IRR (95% CI) | IRR (95% CI) |
| National | Ref | 1.71 (0.77 – 3.81) | 1.43(0.64 – 3.18) | 0.83(0.37 – 1.84) |
| Freetown | Ref | 1.25(0.56 – 2.78) | 0.72(0.32 – 1.61) | 0.57(0.25 – 1.26) |
| Provincial | Ref | 2.07(0.93 – 4.60) | 1.97(0.89 – 4.40) | 1.03(0.46 – 2.29) |

IRR=incident rate ratio

### Regional and periodic changes in patient referrals in Sierra Leone

Figure 4 shows the national median monthly referrals disaggregated by region over time. The National Referral System started gradually following its launch in the post-Ebola period and was augmented by the rollout of the ambulance system. The referral system was maintained after the introduction of NEMS but declined during and after the COVID-19 pandemic. The decrease in patient referrals during COVID-19 was more pronounced in provincial hospitals than those in Freetown. Likewise, the ambulance system contributes more to the provincial referrals than referrals in Freetown (Figure 4).

**Figure 4:**
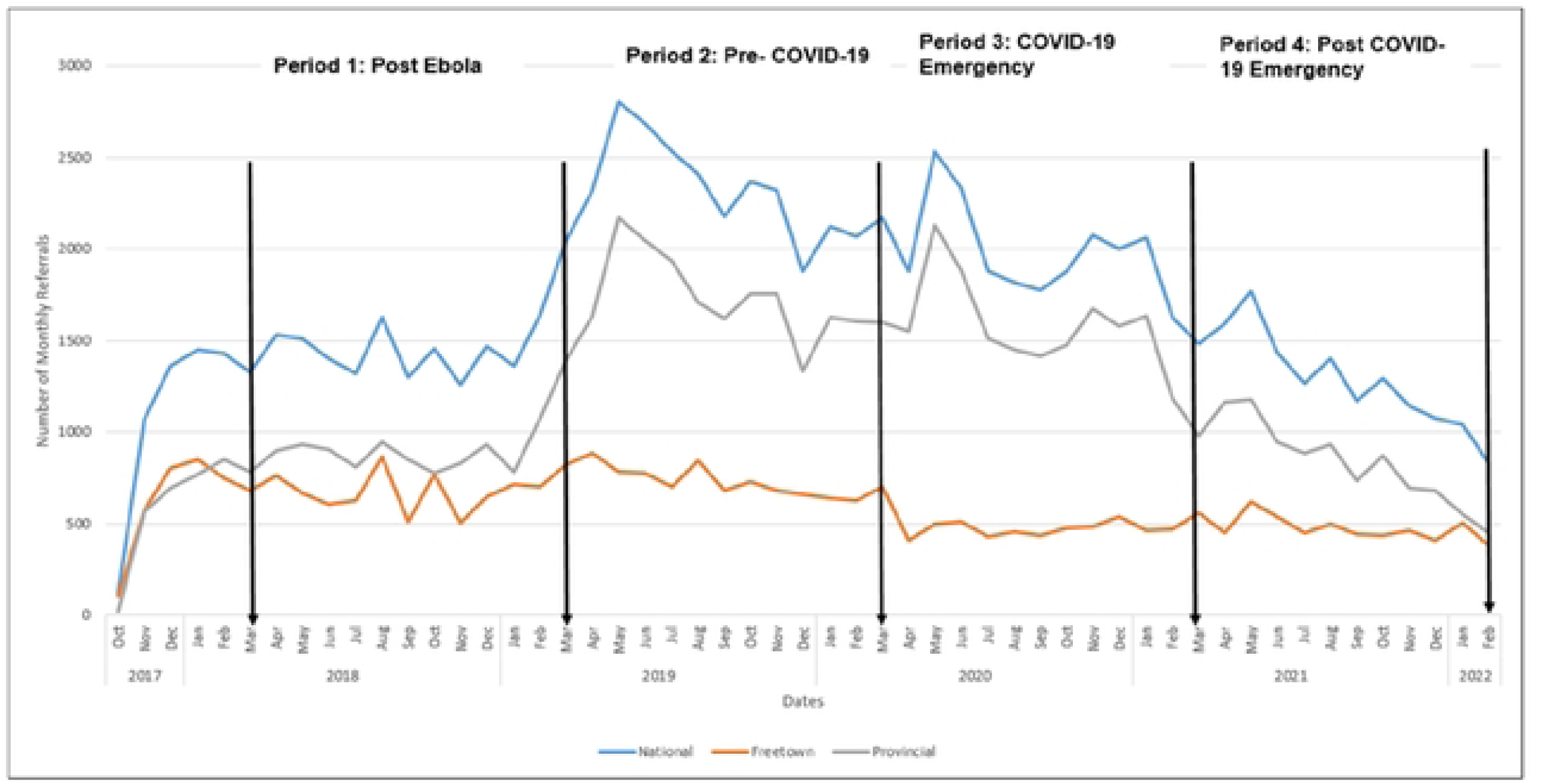
National monthly referrals by region and across periods.

### National monthly bed occupancy across regions and periods

After the establishment of the national referral coordination system, the monthly bed occupancy rate has initially improved. The monthly bed occupancy rate dropped from 730 before the epidemic to 220 during the epidemic. The decline in bed occupancy was more pronounced in Freetown than in provincial regions (Figure 5).

**Figure 5:**
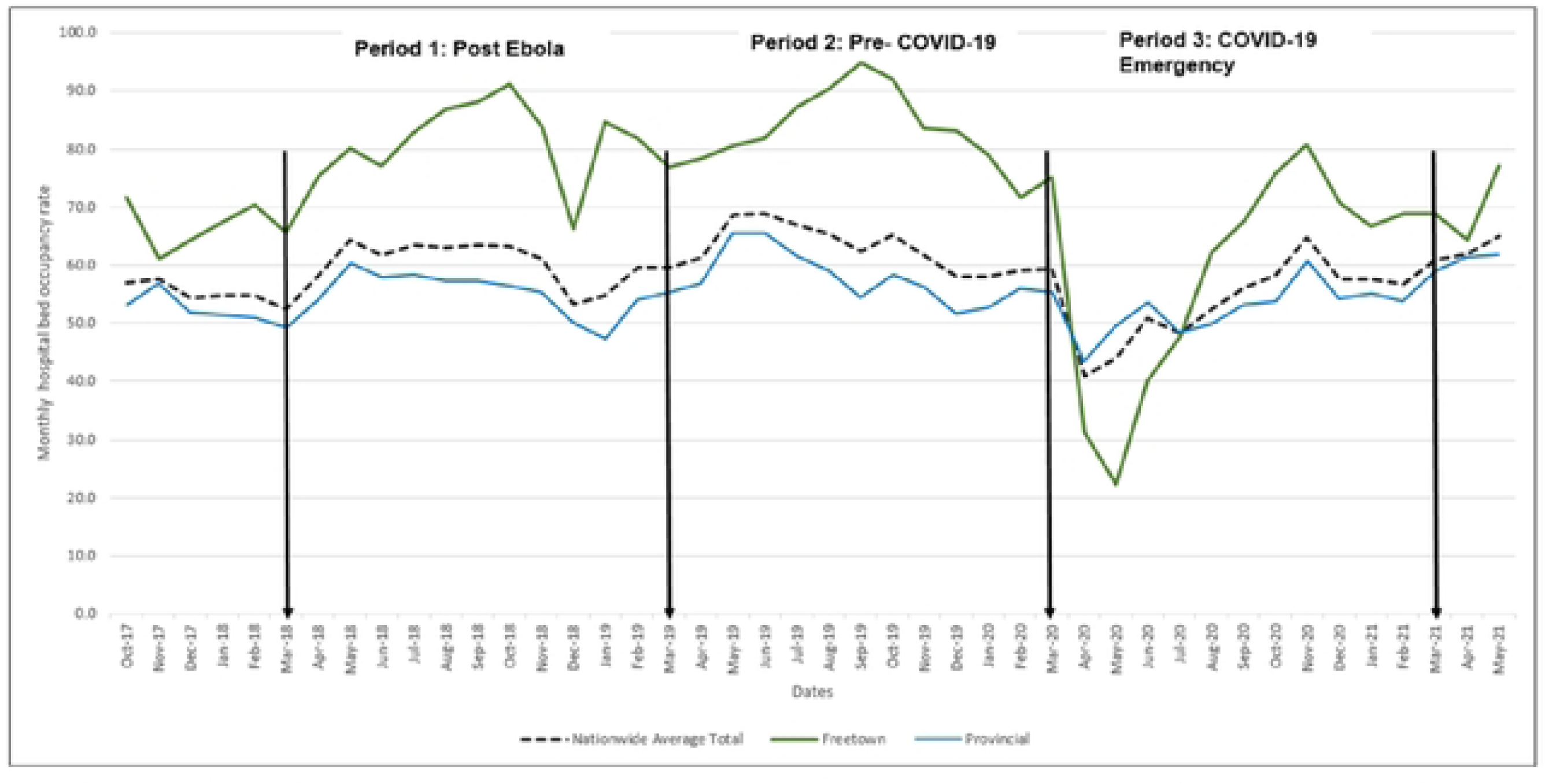
National monthly hospital bed occupancy rates by region across periods.

## DISCUSSION

We describe the National Referral System established in 2017 and examine the effect of COVID-19 on hospital bed occupancy and the use of the National Referral System [9]. Our evaluation of this system, shaped by lessons learned from the 2014-2016 Ebola outbreak, highlights the health system’s resilience in Sierra Leone in the aftermath of a large-scale epidemic [23].

Our findings demonstrated 3 key points. First, the feasibility of establishing a national referral system for post-public health emergencies. Second, the essential role of the National Ambulance System in enhancing service accessibility both for essential and emergency services in the COVID-19 pandemic. And third, the differential impact of COVID-19 on patient referrals and hospital bed utilization between provinces and Freetown. These findings are expected to provide actionable recommendations that will protect essential health services in future public health emergencies.

One of our key findings was that immediately after the referral coordination system was established in 2017, there was a gradual increase in the number of referrals and bed occupancy. Establishing a national referral system reflects the focus of the Sierra Leone National Ebola Recovery Strategy in restoring and strengthening access to basic health services [24]. Sierra Leone’s third National Health Sector Strategic Plan (2021–2025) included interventions to provide health services to all and achieve universal health coverage by 2030 [25]. A strong national referral system is essential to achieving this goal, which further highlights the need to strengthen national referral services in Sierra Leone.

We found geographic variation in referrals and bed utilization during the pandemic. The monthly referrals in Freetown, Sierra Leone’s capital, during the COVID-19 pandemic were less affected than provincial referrals. This could be due to infrastructure challenges facing provincial areas, or the centralisation of services in the capital, or perhaps the repurposing of ambulances to support the COVID-19 referral system in Freetown. A 2016 systematic review highlighted the value of decentralization of services in addressing equity issues, which is necessary to mitigate the impact of public health emergencies on referrals to remote settings [26]. Paradoxically, our study shows that the impact of the pandemic on hospital bed utilization is more pronounced in the capital city than in the province. The capital city has more resources to maintain adequate hospital bed utilization in pandemics, but closure of key hospitals due to outbreaks of nosocomial COVID-19 infections between April and June 2020 may have affected the bed occupancy. Notably, the national children’s hospital in the capital Freetown was closed in the early part of the pandemic. This is an important lesson that we should learn from and not repeat the same mistakes. In a reasonable and well-functioning emergency preparedness and response system, hospital services should not be shut down.

Drawn from the experience of the 2014-2016 Ebola Virus Disease epidemic, several mitigation measures were implemented during COVID-19 to protect essential health services [27]. The Emergency Operations Centre was reactivated early in the pandemic to develop and implement policies to safeguard essential health services and to restructure the health systems, including the national ambulance service [28]. These mitigation measures were planned to build a resilient referral system. However, the referral system did not recover after the COVID-19 disruptions as the reductions in the number of referrals remained low right through the pandemic. This persistently low numbers of referral could be due to the dwindling donor support for the national ambulance system during the funding transition in March to June 2024. Therefore, sustainable investment and mainstreaming of activities are critical steps to build resilient and sustainable health systems in Sierra Leone [29].

Our study has limitations. The reduction in hospital bed utilisation may be due to changes in health-seeking behaviour following the COVID-19 pandemic. It may not reflect unavailability of services Second, we utilized secondary data to assess referrals and hospital bed utilization. Although there are referral coordinators in all the 18 hospitals included in this study, they may have missed some data on bed occupancy and patient referral. We excluded hospitals in Western Rural of Sierra Leone as their data is not part of the national referral system. Finally, the 95% confidence intervals for the monthly referral trends are wide. Therefore, we were unable to state any significant differences due to the overlapping confidence intervals. Nevertheless, our study is the first to describe the feasibility of establishing the national referral system in Sierra Leone and the differential impact of the COVID-19 on this system. Our findings have several implications for a resilient and sustained health system in low-income countries such as Sierra Leone.

## Conclusion

Findings demonstrate that it is feasible for resource-limited countries such as Sierra Leone, to set up national referral systems to enhance access to services following public health emergencies. The impact of COVID-19 on patient referrals and bed utilisation varied across provincial regions and the capital, Freetown. Finally, planned mitigation measures to build a resilient referral system were not restored following the COVID-19 outbreak, as the reduction in referrals remained low throughout the pandemic due to reduced donor support. Local sustainable investments and mainstreaming of activities are key steps towards building a resilient and sustainable health system in Sierra Leone.

## Acknowledgements

SM is supported by a Tier 2 Canada Research Chair in Mathematical Modeling and Program Science.

## Funding

This study was conducted with support from the Canadian Institute of Health Research: (grant number: CIHR: WI1-179883).

## Conflict of interest

None to declare

## Author’s contribution

Conceptualization and study design: DY, IFM, MSK, AKC, SS, SM, SoS, and SL

Data curation and validation: DY, BM, SS, SoS, JSK, SM, HKL,TMB, MK and FM

Methodology and formal analysis: HKL, SMK, SoS, ACK, MJV, FM, MK and JSK

Funding acquisition, resources, supervision: MV, SK, SS, SL, SM, SMK, AKC

Writing – original draft preparation: MTS,SL, SS, SoS, ACK and DY

Writing – review & editing: DY, SL, ACK, SoS, ARW, MJV, SoK, BM, MK, HNK, FM, SK, and SM

## Availability of data

The datasets (doi: 10.6084/m9.figshare.28417283) used and/or analyzed for this study are available from: https://figshare.com/account/items/28417283/edit.

## Notes

### Competing Interest Statement

The authors have declared no competing interest.

### Author Declarations

Ethics approval was obtained from the Sierra Leone Ethics and Scientific Review Committee of the Ministry of Health, Government of Sierra Leone with ethical approval number 020/05/2023. The study involved secondary analysis of anonymised service level data, abstracted under a waiver of informed consent from the ethics committee with the approval of National Emergency Medical Services (NEMS)

